# Community-Driven Copy Number Variant Discovery at Scale: Results from a Rare Disease Genomics Hackathon

**DOI:** 10.1101/2025.08.08.25333317

**Authors:** Ming Yin Lun, Jennifer E. Posey, Jesse D. Bengtsson, Haowei Du, Rituparna Sinha Roy, Lei Yang, Sebastian Ochoa, Bo Yuan, Maddie Gillentine, Anna Lindstrand, Claudia M. B. Carvalho

## Abstract

**Purpose:** Copy number variants (CNVs) are a major contributor to rare genetic diseases, but their detection and interpretation from short-read genome sequencing (srGS) data remain challenging, especially at scale. Large amounts of existing srGS data remain under-analyzed for clinically relevant CNVs.

**Methods:** During a collaborative Hackathon, we developed and applied scalable CNV analysis workflows to srGS data from three unsolved, exome-negative, rare disease cohorts: Primary Immunodeficiency (N = 39), Turkish developmental disorders (N = 31), and data from the Genomics Research to Elucidate the Genetics of Rare diseases (GREGoR) (N = 1437). We employed Parliament2 for structural variant (SV) calling, Mosdepth and SLMSuite for read-depth–based quality control and CNV detection, and R Shiny-based visualization tools. We also constructed an SV/CNV variant database with population frequency and pathogenicity annotations, applied DBSCAN clustering for internal allele frequency estimation, and used a 3-way annotation strategy to aid interpretation.

**Results:** Our pipelines identified high-confidence CNVs and streamlined interpretation across cohorts. Within 2 days, the Hackathon yielded 39 candidate pathogenic SVs. The tools and workflows enabled rapid filtering, prioritization, and visualization of clinically relevant variants.

**Conclusion:** This community-driven effort demonstrates the feasibility and utility of scalable CNV analysis for accelerating diagnosis and discovery in rare disease cohorts using srGS data.

## Introduction

The last two decades have witnessed transformative advances in rare disease genomics, driven by collaborative efforts such as the Centers for Mendelian Genomics (CMG), GREGoR,^1^and international platforms like the Matchmaker Exchange^2^. These programs have collectively analyzed tens of thousands of families, greatly expanding our understanding of gene-disease relationships. However, with this scale comes a persistent challenge: maximizing the diagnostic and discovery potential of existing srGS datasets. Among the most underutilized resources in this context are CNVs, which remain difficult to detect and interpret from srGS data due to issues such as non-uniform coverage, mapping biases, and high false positive variant call rates.

Despite the availability of CNV calling tools for srGS data, their integration into diagnostic workflows has been inconsistent, and the scalability of CNV interpretation remains a bottleneck. As the field transitions into a new era of long-read sequencing and multi-omic integration, it is critical not to overlook the chromosomal variation that remains hidden in legacy datasets. Efficient, scalable, and interpretable CNV workflows are needed to bridge this gap.

In response, we organized a multi-institutional Hackathon with the overarching goal of implementing robust CNV analysis pipelines tailored to large unsolved rare disease cohorts. The structure of the Hackathon was built upon prior experiences and was inspired by previously published frameworks for collaborative rare disease genomics ^3–5^. We emphasized a dual goal of analysis and education—training participants in the implementation of CNV workflows while generating interpretable results in parallel. The primary resources included an internally curated and annotated CNV/SV database, along with visualization platforms such as IGV and VizCNV, a CNV visualization tool, integrated into the user interface^6, 7^. Participants were divided into 3 multidisciplinary teams; each assigned a unique cohort for analysis. The teams are structured such that each team has a disease expert, a bioinformatician, and a genomic analysist expert. This structure enabled both community-based learning and scalable CNV discovery, advancing our shared goal of leveraging short-read sequencing to its fullest potential in rare disease genomics.

## Materials and Methods

### SV calling

Parliament2 was used for SV calling^8^. The latest version (0.1.11) of Parliament2 was run inside a docker container using the following command ‘docker run dnanexus/parliament2:latest –bam input.bam –bai input.bam.bai -r ref.fa –fai ref.fa.fai –breakdancer –breakseq –manta –lumpy -- delly_deletion --delly_insertion --delly_inversion –genotype’.

### Generating read depth segments, CNV calls, and plots

BAM files were generated by aligning Illumina short-read sequencing data to the human reference genome, with GRCh37 used for the PID and TDD cohorts, and GRCh38 used for the GREGoR cohort. BAM files are then processed using Mosdepth 0.3.7 (-Q 30 -b1000 -x -n) ^9^to generate BED files, which store genomic positions and corresponding coverage. The log_2_ ratio of the coverage datapoints were segmented using SLM algorithm in the SLMSuite^10^. Log_2_ ratio is calculated as follows:

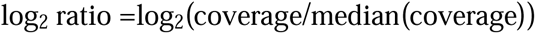

All qualified segments, those larger than 10 kb and with log_2_ ratio >= −2, are extracted. Small segments are excluded because of the alignment challenges caused by small genomic repeats. The qualified segments are used to generate CNV calls and log_2_ ratio read depth plots in R using ggplot2. For more details, please refer to vizCNV^7^.

### Read depth quality control (QC)

Qualified segments in autosomes for each subject are used to calculate the standard deviation (SD) of log_2_ ratio which we use to evaluate the spread of the read depth signal across the genome. This QC metric was developed to emulate the derivative log ratio (DLR) spread used to control signal to noise in array comparative genomic hybridization (aCGH)^11^. Based on the distribution, SD of 0.38 was used as a cutoff, where any value higher or equal to 0.38 will be deemed as an outlier. (Figure 1A, red dotted line). There were 0, 1, and 34 samples in the TDD, PID, and GREGoR cohort, respectively, with SD higher than cutoff. Those samples were removed from further analyses of the study. The correlation between total qualified segment counts and the SD for GREGoR samples showed a negative correlation with R-coeff of −0.835 (Figure 1B). Similar results are shown for the PID and Turkish cohort (Supp Figure 1). The difference between a sample below the cutoff and above the cutoff can be seen in Figure 1C and 1D.

**Figure 1:**
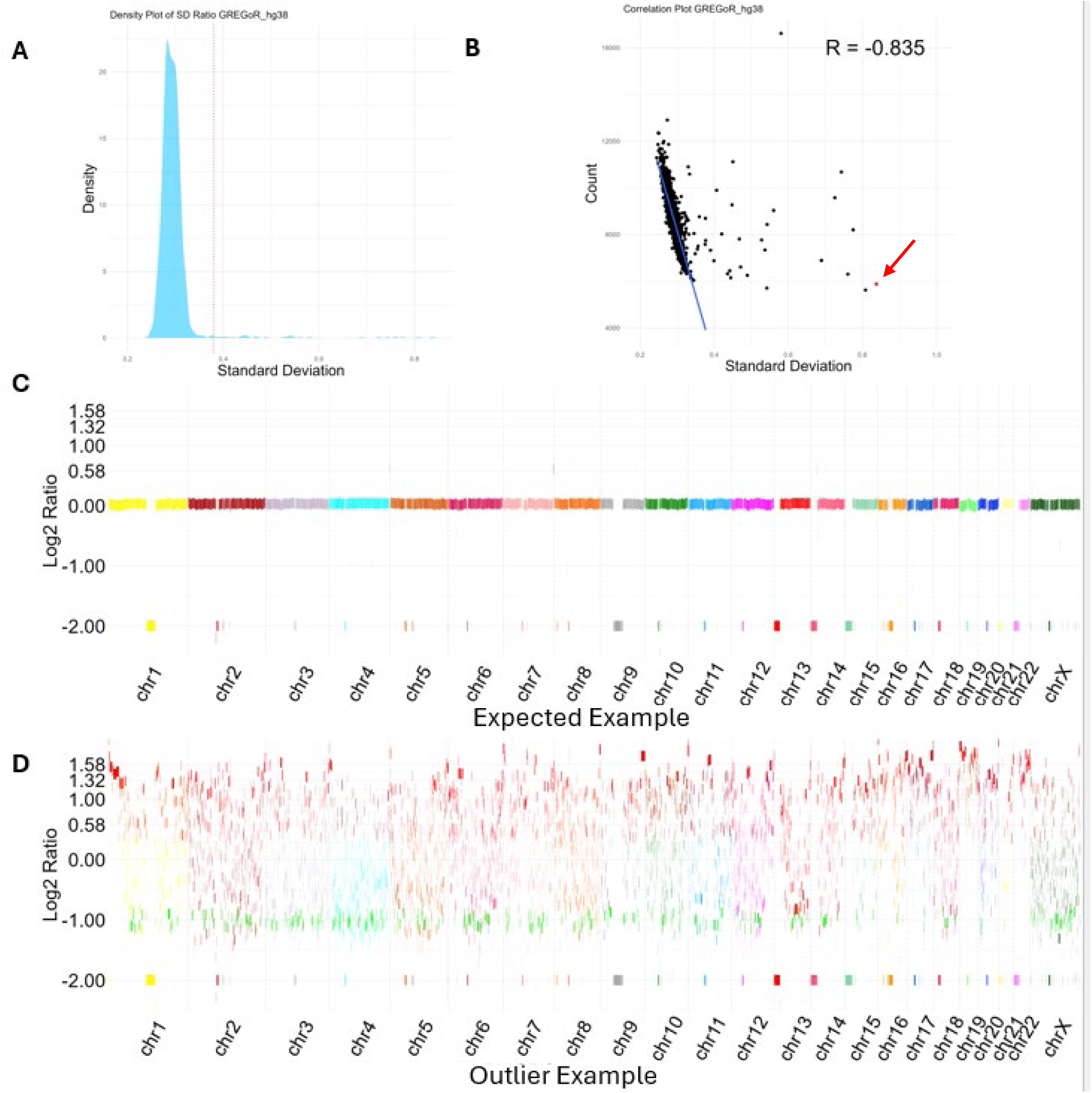
GREGoR cohort read depth QC results. A) Density plot of standard deviation of log2 ratio for all qualified segments in N = 1,420 samples. Red dotted line shows the log2 ratio standard deviation (SD) threshold of 0.38. B) Correlation plot between qualified segments counts per sample and corresponding sample SD value. The negative correlation suggests that higher SD could lead to lower CNV resolution and higher rates of false-negative CNVs. C) Typical log2 ratio graph displaying genome-wide distribution per chromosome of a sample with SD < 0.38. Notice the consistent long line in the middle. D) Representative genome-wide distribution per chromosome of the log2 ratio from one out of 34 outlier samples with log2 ratio SD >= 0.38.

### Clustering and internal allele frequencies

DBSCAN was used to cluster CNV/SV calls. CNV/SV calls are first grouped by type (DEL, DUP, INV, BND), the left and right breakpoints are then represented on a cartesian graph on x and y axis, respectively, and clustered using DBSCAN (epsilon=500, min_samples=2) ^12^. This allows the clustering of not only calls on the same chromosome but also calls with breakpoints in two different chromosomes (e.g. translocations). The internal allele frequency can be calculated based on the number of calls in each cluster. (Figure 2A)

**Figure 2.**
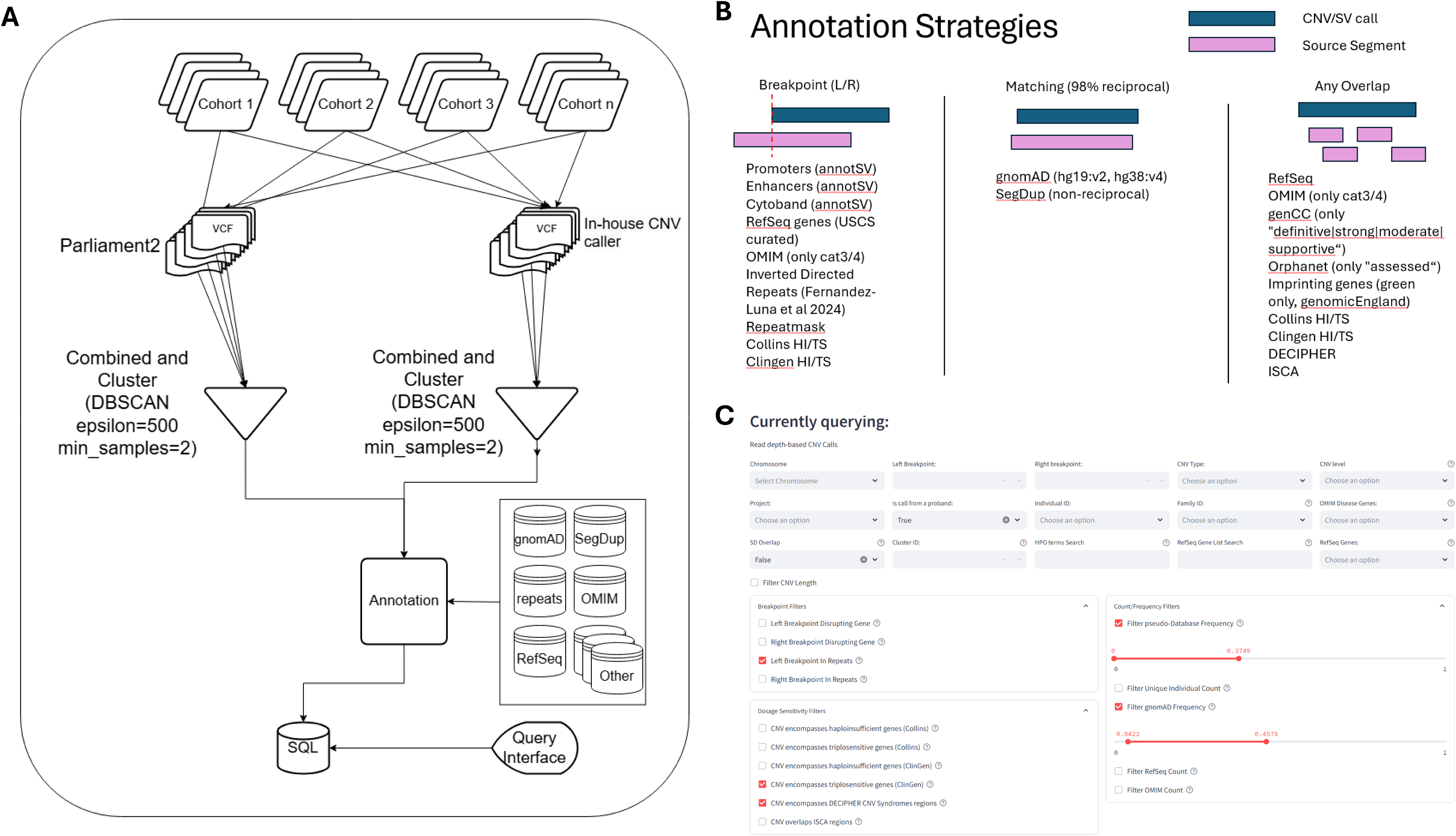
CNV/SV Processing, Annotation Strategies, and Interactive Query Interface. A) Overview of processing pipeline. All cohorts are processed by Parliament2 and an in-house CNV caller. The variant calls are then clustered using DBSCAN and annotated using BEDanno. The data is then stored in SQL in the backend and queried using streamlit as the frontend. B) CNV/SV annotation was performed based on three different strategies. *Breakpoint:* left and right coordinates of a call; *Matching*: 98% reciprocal overlap; *Any Overlap*: overlap with any percentage without reciprocity. C) User interface for CNV/SV querying using various filters. The interface allows users to filter CNV/SV calls based on various parameters, including chromosome, CNV/SV type, individual and family IDs, and gene lists (OMIM, RefSeq). Breakpoint filters include gene disruption and repeat overlaps. Dosage sensitivity filters incorporate haploinsufficiency and triplosensitivity predictions from Collins and ClinGen, as well as DECIPHER and ISCA regions. Frequency filters enable users to set thresholds based on pseudo-database frequency, gnomAD frequency, and individual counts, facilitating the identification of potentially pathogenic CNVs.

### CNV/SV Annotation

The annotation was performed using sources from both GRCh37.p13 and GRCh38. Three annotation strategies were employed: *Breakpoint*, *Matching*, and *Any Overlap* (Figure 2B). bedtools v2.29.1 were used to facilitate the annotation process. All annotation sources are shown in Table 1^13^.

**Table 1.** Annotation sources and references.

| Annotation | Filtering | Source | Reference |
| --- | --- | --- | --- |
| **Promoters** | NA | AnnotSV | Geoffroy et al 2018<sup>14</sup> |
| **Enhancers** | NA | AnnotSV | Geoffroy et al 2018<sup>14</sup> |
| **Cytobands** | NA | AnnotSV | Geoffroy et al 2018<sup>14</sup> |
| **RefSeq** | NCBI RefSeq curated | UCSC | Pruitt et al 2014<sup>15</sup> |
| **SegDup** | bedtools merge -d 1000,<br>removed PAR regions | UCSC | Bailey et al 2022<sup>16</sup> |
| **RepeatMasker** | NA | UCSC | www.repeatmasker.org/ |
| **clinGen HI/TS** | Score > 3 | UCSC | Rehm et al 2015<sup>17</sup> |
| **Collins HI/TS** | HI $\geq 0.86$, TS $\geq 0.94$ | UCSC | Collins et al 2022<sup>18</sup> |
| **genCC** | Definitive\|strong\|moderate\|supportive | UCSC | DiStefano et al 2022<sup>19</sup> |
| **Orphanet** | “Assessed” only | UCSC | Pavan et al 2017<sup>20</sup> |
| **ISCA** | NA | UCSC | Rehm et al 2015<sup>17</sup> |
| **gnomAD** | NA | gnomAD (GRCh37:v2,<br>GRCh38:v4) | Chen et al 2024<sup>21</sup> |
| **OMIM** | Category 3 and 4 only | OMIM | Amberger et al 2009<sup>22</sup> |
| **DECIPHER** | NA | DECIPHER website | https://www.deciphergenomics.org/ |
| **Imprinting genes** | Green category only | Genomics England | Tucci et al 2019<sup>23</sup> |
| **Inverted and Direct Repeats** | NA | Fernandez-Luna et al 2024 | Fernandez-Luna et al 2025<sup>24</sup> |

For *Breakpoint* annotations, left and right breakpoints as determined by the CNV/SV caller are annotated if any of the source segments encompass the location of the break. For *Matching* annotations, CNV/SV calls are annotated if there is more than 98% reciprocal matching between the CNV/SV call and the source segment (bedtools intersect -f 0.98 -r). Since sequence alignment and variant detection are not always perfect and give slight variability around breakpoints, a reciprocal 98% overlapping parameter ensures that variants can be matched even if they do not have exact breakpoint resolutions. For *Any Overlap* annotation, all source segments that are overlapping with the CNV/SV call are reported, regardless of overlap percentage or reciprocity.

These annotation strategies are catered to retrieve different types of information for each call. *Breakpoint* annotation is important for knowing if a certain gene, promoter, or enhancer is disrupted, and provides information of whether a repeat element maps at the breakpoint. Such information is especially important for inversions and translocations, as copy number neutral SVs often require breakpoint information to derive potential functional consequences of the SV. *Matching* annotation provides population frequency from public databases such as gnomAD. The SegDup source was simplified by merging any segmental duplications that are less than 1 kb apart (bedtools merge -d 1000). The PAR regions were removed from the SegDup track. SegDup annotation also does not require reciprocity as the focus is to determine if the call is 98% overlapping with any SegDup. *Any Overlap* annotation focuses on functional annotation, looking for genic or non-genic regions that are known to cause diseases, for example OMIM genes, Orphanet genes, ISCA regions, DECIPHER regions, etc. (Table 1)^14–20, 22–25^.

### Variant Querying

The user interface was built using python for CNV/SV querying using various filters. The interface allows users to filter CNV/SV calls based on various parameters, including chromosome, CNV/SV type, individual and family IDs, and gene lists (OMIM, RefSeq). Breakpoint filters include gene disruption and repeat overlaps. Dosage sensitivity filters incorporate haploinsufficiency and triplosensitivity predictions from Collins et al. ^18^ and ClinGen, as well as DECIPHER and ISCA regions. Frequency filters enable users to set thresholds based on pseudo-database frequency, gnomAD frequency, and individual counts, facilitating the identification of potentially pathogenic CNVs. (Figure 2C)

## Results

The study analyzed exome-negative probands from three cohorts using short-read genome sequencing with tailored CNV prioritization strategies. Many of the probands were recruited as part of the CMG program.^26^ The PID cohort applied two main strategies: (1) identification of *de novo* CNVs and (2) CNVs overlapping known PID genes defined by the IUIS, excluding variants with poor phenotype correlation or located in repetitive regions. The TDD cohort (35 probands), with known consanguinity population, employed two filtering strategies: (1) prioritizing homozygous CNVs and (2) CNVs overlapping OMIM genes. The GREGoR cohort (1,437 individuals) used a dual strategy: (1) identifying rare CNVs in probands regardless of family structure and (2) detecting recurrent CNVs overlapping dosage-sensitive regions from ISCA, DECIPHER, and ClinGen. CNVs were reviewed using VizCNV to assess log ratio consistency, B-allele frequency, and inheritance. Collectively, these cohort-specific strategies highlight the importance of phenotype-driven and context-aware CNV prioritization in uncovering pathogenic variants in rare disease cohorts.

### Primary Immunodeficiency (PID) cohort

The Primary Immunodeficiency (PID) cohort consisted of 39 trios (probands and parents), where the probands included 24 males, 14 females, and one individual of unspecified biological sex. All participants presented with phenotypic features consistent with childhood-onset immunological disorders. The most frequently observed clinical categories included immune dysregulation— manifesting as complex autoimmunity and recurrent infection, periodic fever syndromes, and severe humoral immune deficiencies, such as common variable immunodeficiency (CVID) and agammaglobulinemia.(Figure 3A)

**Figure 3.**
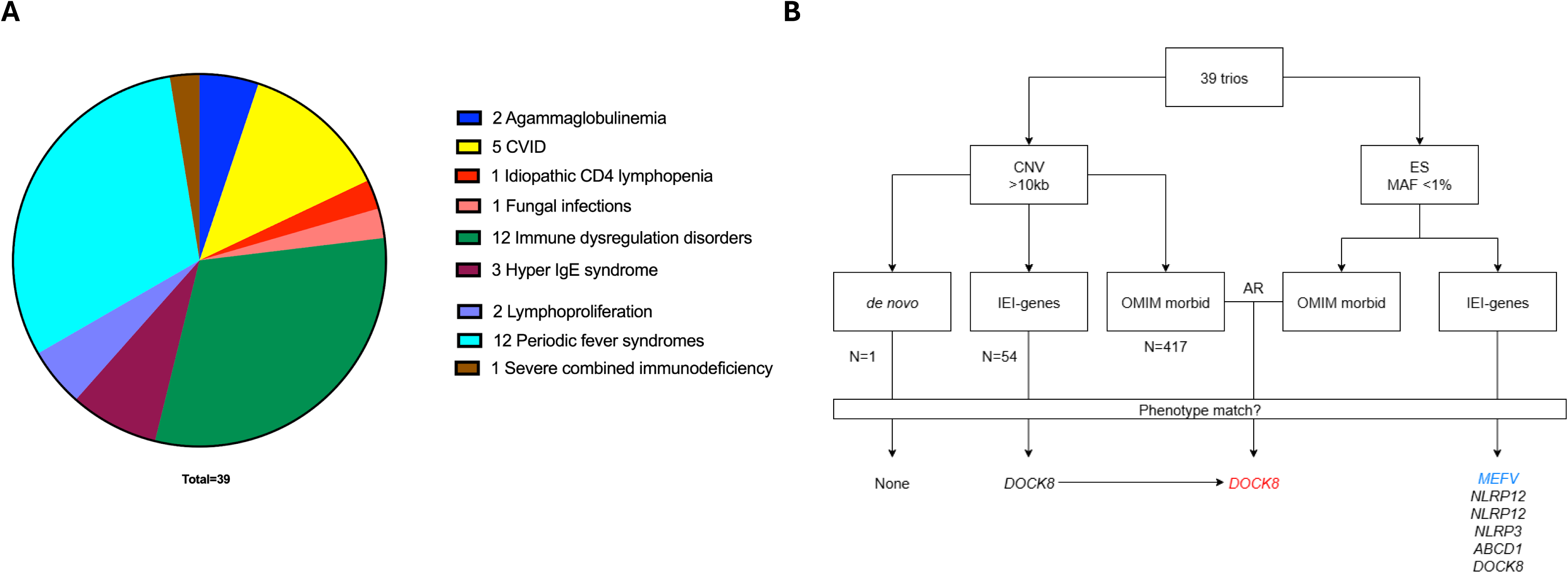
PID cohort overview. A) Main phenotype groups for the PID cohort. The majority of the participants fall into one of two groups: immune dysregulation disorders or periodic fever syndromes. B) The analytical pipeline for the inborn errors of immunity (IEI) cohort is depicted. Copy number variant (CNV), exome sequencing (ES), inborn errors of immunity (IEI), minor allele frequency (MAF), and Online Mendelian Inheritance in Man (OMIM).

To identify candidate variants, both single nucleotide variants (SNV)/indel analysis from exome sequencing (ES) and CNV analysis from srGS were performed independently and in combination (Figure 3B). The CNV analysis followed two strategies: (1) the identification of any *de novo* CNV, and (2) the identification of CNVs overlapping genes previously associated with PID, as defined by the International Union of Immunological Societies (IUIS)^27^. For both strategies, CNVs larger than 10 kb were considered, and those mostly overlapping segmental duplications were excluded. CNVs found in PID-related genes were further assessed for phenotype correlation, and those showing complete mismatch with the participant’s clinical presentation were excluded.

In the *de novo* CNV analysis, one notable finding was a 59 kb deletion encompassing a long noncoding RNA antisense to *CSMD1* (HGNC:14026). However, this CNV was not consistent with the participant’s phenotype and was excluded. Filtering for CNVs in PID-related genes identified seven CNVs within the cohort. Of these, four overlapped genes frequently affected by common repetitive elements—*CFHR3* (HGNC:16980), *CFHR1* (HGNC:4888), *CFHR4* (HGNC:16979), and *CSF2RA* (HGNC:2435)—and were present across multiple participants, suggesting limited specificity. Two additional CNVs, involving *NCF1* (HGNC:7660) and *CD59* (HGNC:1689), did not align with the participants’ clinical features and were also excluded. This process resulted in the identification of a single strong candidate CNV: a deletion involving *DOCK8* (HGNC:19191) (chr9:298,000–311,000) in participant P11.

Re-analysis of ES data involved filtering for variants with a minor allele frequency (MAF) <1% in gnomAD v2, focusing on OMIM-morbid genes, and assessing genotype-phenotype concordance. One participant had a clinical diagnosis of adrenoleukodystrophy, supported by a pathogenic *ABCD1* (HGNC:61) variant that had recently been reclassified from a variant of uncertain significance (VUS) to pathogenic. In another case, a variant in *MEFV* (HGNC:6998)— a gene associated with autosomal dominant and recessive forms of familial Mediterranean fever—was identified, though the variant’s classification remained conflicting in ClinVar. Additionally, five heterozygous candidate VUSs were identified across the cohort (Table 2, Figure 3B).

**Table 2.** Summary of pathogenic or likely pathogenic SNVs/CNVs identified for PID cohort.

| Part | Phenotype | Variant(s) | Zygoty | ClinVar | OMIM # | Inheritance | MAF gnomAD ES | Contribution to phen |
| --- | --- | --- | --- | --- | --- | --- | --- | --- |
| P11 | HIES | c.5599 T>G<br>HET_DEL<br>(*DOCK8*,<br>chr9:298000-311000) | Comp Het | Absent | *DOCK8*<br>Deficiency (HIES)<br>#243700 | AR | 0<br>n/a | Yes |
| P14 | HIES | c.4283 C>A | Het | VUS | *DOCK8*<br>Deficiency (HIES)<br># 243700 | AR | 0.0002 | No |
| P25 | Periodic<br>Fever | c.2084 A>G | Het | Likely<br>pathogenic | *MEFV*<br>*608107 | AD/AR | 0.0036 | Likely |
| P31 | Periodic<br>Fever | c.1339 C>G | Het | VUS | *NLRP12*<br>*609648 | AD | 0.0001 | No |
|  |  | c.608 C>T | Het | VUS | *NLRP3*<br>*606416 | AD | 0.0001<br>8 | No |
| P39 | HLH | c.1330 C>G | Het | VUS | *NLRP12*<br>*609648 | AD | 0 | No |

Part: Participant; Het: Heterozygous; DEL: Deletion; VUS: Variant of unknown significance; HIES: Hyper-IgE syndrome 1; HLH: Hemophagocytic lymphohistiocytosis; AD: Autosomal dominant; AR: Autosomal recessive; Phen: Phenotype

The final analysis employed an integrative strategy combining any rare CNV with any rare variant (MAF <1%) in genes associated with autosomal recessive inheritance. This approach led to the identification of compound heterozygous variants in *DOCK8* in participant P11, who exhibited a clinical phenotype consistent with AR DOCK deficiency^28^. The two *DOCK8* variants were confirmed to be in trans and were validated by Sanger sequencing (Table 2, Figure 3B).

### Turkish Development Delay (TDD) cohort

The Turkish cohort consisted of 35 individuals of Turkish descent, with a high rate of reported parental consanguinity^29^. The gender distribution was balanced, with 54% females and 46% males. Clinical annotation revealed a range of phenotypic complexity, with individuals presenting between 1 and 11 phenotype terms (median: 3). The most prevalent clinical presentations included arthrogryposis (n=7), Klippel-Feil syndrome (n=2), and hypergonadotropic hypogonadism (n=2). Additional unique diagnoses included Moebius syndrome, sensorineural hearing loss, PEHO syndrome, dwarfism, Goldenhar syndrome, and isolated nystagmus, each represented by a single individual. (Figure 4A/B, Supp Figure 2)

**Figure 4.**
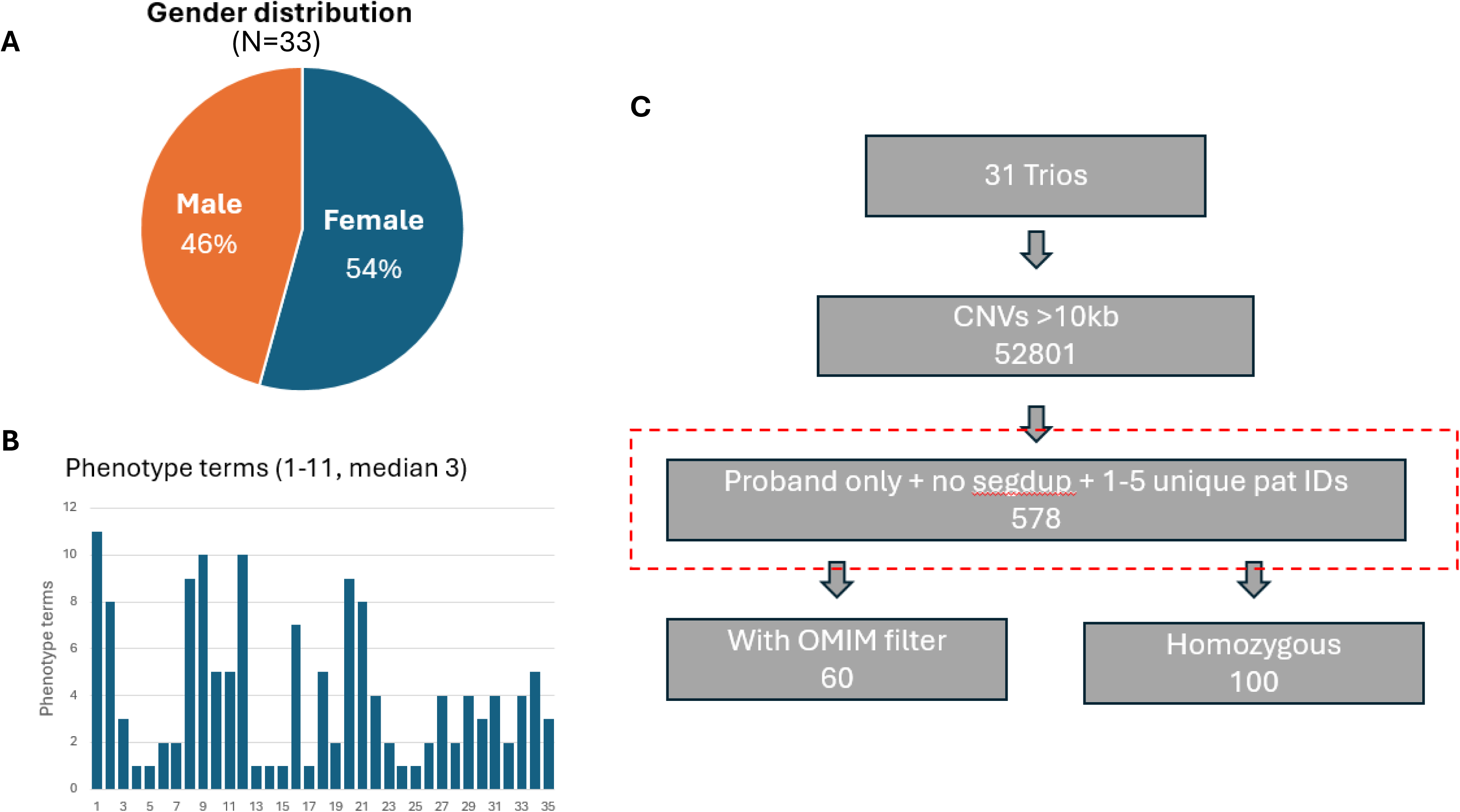
Turkish Developmental Disorder cohort overview. A) Gender distribution. B) Phenotype term distribution. C) CNV analysis pipeline.

Structural variant analysis of the 31 trios identified a total of 52,801 CNVs larger than 10 kb. We prioritized 578 CNVs that were observed only in probands, did not overlap segmental duplications, and clustered with ≤5 unique participant IDs. From this subset, we applied two refinement strategies: (1) filtering based on OMIM gene overlap, which yielded 60 CNVs, and (2) selecting homozygous CNVs, resulting in 100 candidates for further investigation. (Figure 4C)

Upon review, four clinically relevant CNVs (three pathogenic and one variant of unknown significance, VUS) were identified in four individuals with neurodevelopmental and/or congenital phenotypes. The clinical features, genomic coordinates, inheritance, and molecular diagnoses are summarized below.

BH9683-1 is a male presenting with developmental delay, cardiomyopathy, microcephaly, myopathy, and cerebral atrophy. CNV analysis revealed a 3.92 Mb deletion at 15q11–q13 (chr15:23,754,000–28,437,000). The deletion was paternally inherited, confirming a molecular diagnosis of Prader-Willi syndrome (MIM#176270) ^30, 31^ (Supp Figure 3).

BH10684-1 is a female clinically diagnosed with PEHO (MIM#260565) (progressive encephalopathy with edema, hypsarrhythmia, and optic atrophy) syndrome, optic nerve atrophy, cerebral atrophy, and severe intellectual disability (IQ 20–35). A 3.27 Mb deletion overlapping the same critical 15q11–q13 region (chr15:23,754,000–28,438,000) was identified and found to be maternally inherited. The inheritance pattern supports a diagnosis of Angelman syndrome ^30^ (Supp Figure 4).

BH13688-1 is a female diagnosed with arthrogryposis. Molecular analysis revealed an 84 kb deletion (chrX:64,159,000–64,243,000) partially encompassing *ZC4H2* (HGNC:24931). This deletion was confirmed to be *de novo*. (Supp Figure 5) Given the clinical presentation and known disease association of *ZC4H2* disruptions with arthrogryposis and Wieacker-Wolff syndrome (MIM#314580), the variant is considered pathogenic ^32, 33^.

BH13674-1 is a female presenting with epilepsy and developmental delay/intellectual disability (DD/ID). A 149 kb intronic deletion was detected within *MACROD2* (HGNC:16126)

(chr20:14,735,000–14,884,000). While the clinical correlation is suggestive, further functional studies or additional cases are needed to determine pathogenicity ^34^(Supp Figure 6).

Genomic findings and phenotype correlation is summarized in Table 3.

**Table 3.** Summary of pathogenic or likely pathogenic CNVs identified for TDD cohort.

| **ID** | **Sex** | **Genomic Coordinates**<br>**srGS (hg19)** | **Size**<br>**(bp)** | **Clinical**<br>**description** | **Genotype** | **Inheritance** | **Contribution to**<br>**phen** |
| --- | --- | --- | --- | --- | --- | --- | --- |
| **BH9683-1** | M | Chr15:<br>23,754,000-28,437,000 | 4,683,000 | Dev delay,<br>Cardiomyopathy,<br>Microcephaly,<br>Myopathy,<br>Cerebral atrophy | 15q11-13<br>deletion | Paternal | Yes |
| **BH10684-1** | F | chr15:23,703,000-28,438,000 | 4,734,000 | PEHO<br>syndrome,<br>Optic nerve atrophy,<br>Cerebral atrophy,<br>Severe IF (IQ 20-35) | 15q11-13<br>deletion | Maternal | Yes |
| **BH13688-1** | F | chrX:64,159,000-64,243,000 | 84,000 | Arthrogryposis | *ZC4H2*<br>partial deletion | *De novo* | Yes |
| **BH13674-1** | F | Chr20:<br>14,735,000-14,884,000 | 149,000 | Epilepsy<br>DD/ID | 150kb<br>intronic deletion<br>in<br>*MACROD2* | Paternal | VUS |

srGS: short-reads genome sequencing; M: Male; F: Female; VUS: Variant of unknown significance; Phen: Phenotype

### GREGoR cohort

The GREGoR cohort consisted of a total of 1,437 individuals. Following the exclusion of data that did not meet the quality control threshold for log□ ratio standard deviation (SD > 0.38; Figure 1), the final cohort included 400 proband-parent trios (n=1,200 individuals), as well as 156 additional families comprising either proband-relative pairs or proband-only structures (n=237 individuals), as detailed in Table 4. The phenotypic spectrum represented within this cohort was consistent with the GREGoR Consortium’s study population^1^, which focuses on families with suspected Mendelian disorders for whom a definitive molecular diagnosis has not yet been established.

**Table 4.** Summary of individuals included in the analysis for GREGoR cohort, categorized by family structure.

| # Individuals | Trio | Not trio | Total |
| --- | --- | --- | --- |
| **Total** | 1200 | 237 | 1437 |
| **Probands** | 400 | 156 | 556 |
| **Parents** | 800 | NA | 800 |
| **Other (parent or other relative)** | NA | 81 | 81 |

Two complementary analysis strategies were employed to optimize the detection of both non-recurrent and recurrent copy number variants (CNVs). The first strategy (“Analysis #1”; Figure 5A) focused on the identification of unique, non-recurrent CNVs present in probands. Given that the cohort included a mixture of family structures, including non-trio configurations, the demonstration of *de novo* status was not required for CNV prioritization under this approach. To reduce the number of false-positive candidate CNVs, variants mostly overlapping segmental duplications were excluded from downstream analysis. This strategy yielded a total of 4,178 candidate CNVs that required additional review and interpretation.

**Figure 5.**
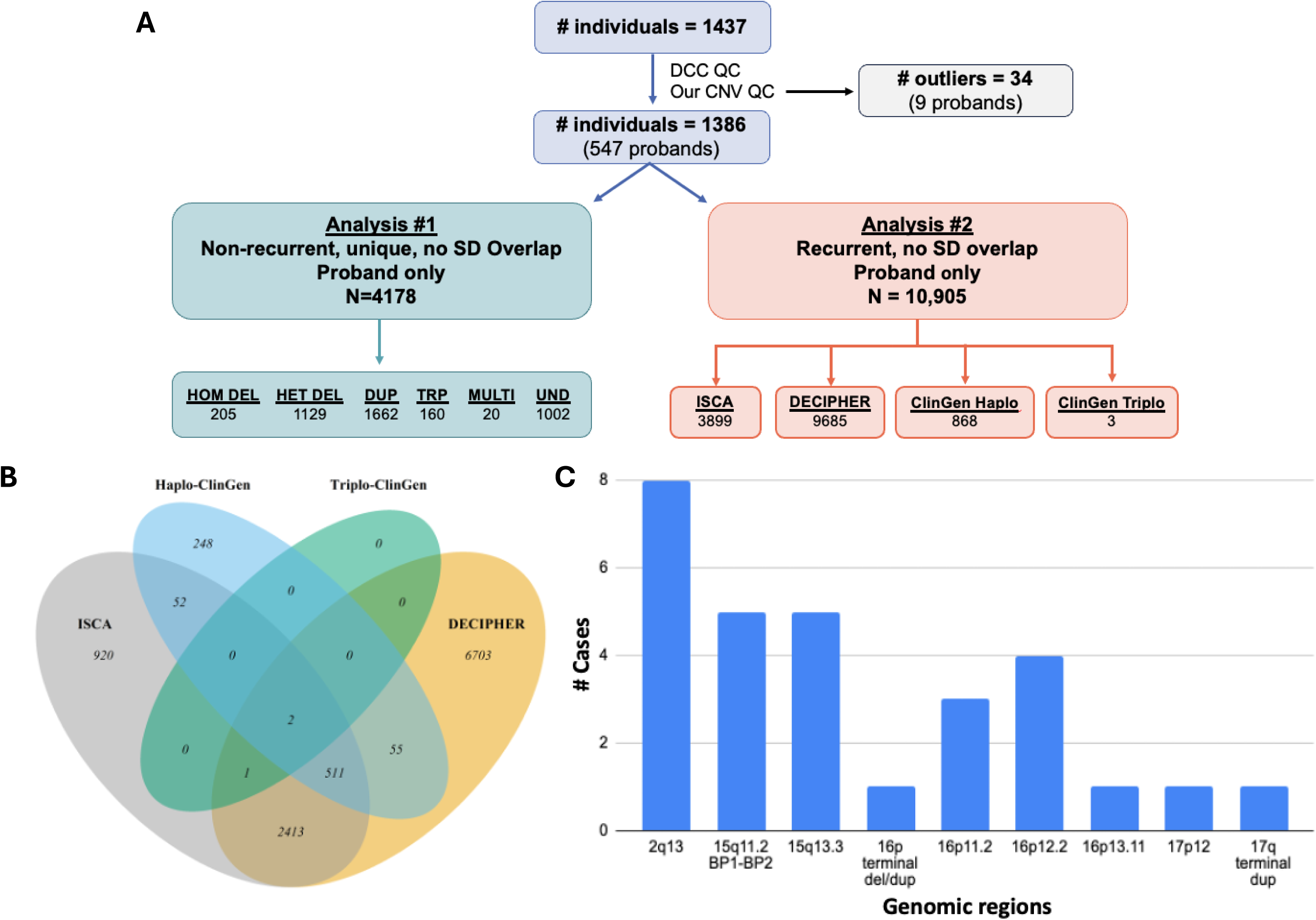
Analytical Framework for CNV Prioritization and Overlap with Dosage-Sensitive Regions. A) Flowchart illustrating the two analytical strategies implemented to identify both non-recurrent (Analysis #1) and recurrent (Analysis #2) copy number variants (CNVs). In Analysis #1, prioritized CNVs are categorized by type: homozygous deletions (HOM DEL), heterozygous deletions (HET DEL), duplications (DUP), triplications (TRP), complex multi-copy variants (MULTI), and variants of undetermined type (UND). In Analysis #2, prioritized CNVs are stratified based on their overlap with dosage-sensitive regions annotated in three reference databases: ISCA, DECIPHER, and ClinGen haploinsufficiency/triplosensitivity scores. B) Venn diagram demonstrating CNVs identified as overlapping with reported dosage sensitive regions from the ISCA, DECIPHER, ClinGen haplosensitivity, and ClinGen triplosensitivity databases. C) Bar chart demonstrating 36 CNV calls each having at least 50% genomic overlap with established ISCA and DECIPHER dosage sensitive regions. The number of probands observed to have each recurrent CNVs (y-axis) is plotted across the indicated genomic regions (x-axis).

The second analysis strategy (Analysis #2”; Figure 5A) applied to the GREGoR dataset was designed to detect recurrent CNVs that overlap dosage-sensitive genomic regions, which were defined based on annotations from three reference databases: ISCA, DECIPHER, and ClinGen haploinsufficiency/triplosensitivity scores. This approach identified 10,905 candidate CNVs requiring further evaluation. To enhance the efficiency of diagnostic review, 36 CNVs demonstrating at least 50% reciprocal overlap with regions designated as dosage sensitive in both ISCA and DECIPHER were prioritized for initial manual review and visualization. (Figure 5B)

Following the prioritization of CNVs from both analysis approaches, we conducted an analysis review of each CNV using VizCNV: Read-depth plots were reviewed for a consistent mean log2 ratio across the CNV region, supporting the identified CNV. Each call was further evaluated for concordance between the CNV prediction and B-allele frequency (BAF) patterns of nearby SNVs. Inheritance status was assessed by examining parental read-depth and BAF plots to determine whether the CNV was inherited or *de novo*.

Additional case information, including family structure, phenotype, and any molecular diagnoses reported within the GREGoR consortium, was also reviewed for each case. The overall numbers of detected CNVs are summarized in Figure 5C. Below, we highlight two representative examples that demonstrate the utility of this approach in a rare disease cohort.

The first case (Supp Figure 7A) involves a trio enrolled due to clinical suspicion of Charcot-Marie-Tooth (CMT) neuropathy, diagnosed clinically in both the proband and her mother. A duplication involving *PMP22* (HGNC:9118) was prioritized for further review. The analysis with VizCNV revealed an increase in the log_2_ ratio overlapping *PMP22* in both the proband and the mother. The B-allele frequency analysis further supported the breakpoint location within a region of segmental duplications, and visual inspection confirmed the maternal inheritance of this pathogenic CNV in the proband ^35^.

The second case (Supp Figure 7B) involves a trio with the proband exhibiting developmental delay, intellectual disability, seizures, and dysmorphic craniofacial features. VizCNV analysis revealed a paternally-inherited *de novo* terminal deletion/duplication encompassing the p arm of chr16. Breakpoint analysis in IGV suggests an inverted duplication formed by fold-back mechanism^36^.

## Discussion

This study demonstrates the utility of GS-based CNV analysis to improve diagnostic yield across diverse rare disease cohorts. By leveraging trio-based pipelines and applying cohort-specific prioritization strategies, we were able to identify both pathogenic and candidate CNVs in multiple individuals, including those that had previously undergone ES without definitive molecular diagnoses. Our findings illustrate both the strengths and limitations of CNV detection in rare disease genetics and provide insight into the application of tailored analytical approaches across heterogeneous populations.

In the PID cohort, which represents a clinically well-characterized set of participants with inborn errors of immunity (IEI), the use of complementary ES and GS analysis was essential for detecting compound heterozygous variants in *DOCK8*—a known IEI-associated gene—in one individual with hyper IgE syndrome (HIES)^37^. This finding, supported by phenotypic congruence and Sanger confirmation of *in-trans* inheritance, exemplifies the diagnostic potential of integrative rare SNV and CNV analysis in autosomal recessive disease contexts. Importantly, the identification and subsequent filtering of CNVs based on IUIS gene overlap and phenotype correlation minimized false positives, particularly in the context of recurrent CNVs affecting genes within repetitive or polymorphic loci (e.g., *CFHR3/1/4, CSF2RA*). This underscores the importance of clinical-genomic concordance in CNV interpretation and highlights the need for careful curation when applying automated pipelines to immunogenetic disorders.

The Turkish cohort provided a valuable lens into the genetic architecture of rare developmental and congenital disorders in a consanguineous population. The detection of homozygous deletions and rare CNVs intersecting OMIM genes was a key feature of the analytical strategy in this group. However, the three identified pathogenic CNVs were not homozygous but rather recurrent *de novo* deletions in the 15q11–q13 region, consistent with Angelman or Prader-Willi syndromes, as well as a *de novo* deletion of *ZC4H2* associated with arthrogryposis. All three CNVs are consistent with participants’ clinical phenotypes. Notably, the detection of an intronic deletion in *MACROD2* in an individual with epilepsy and developmental delay exemplifies the challenges in interpreting variants of uncertain significance and the need for expanded functional validation to resolve their pathogenic potential.

In the GREGoR cohort, which encompassed a broader range of suspected Mendelian disorders, our two-pronged CNV discovery approach was instrumental in managing the scale and diversity of the dataset. The first analysis strategy effectively identified non-recurrent CNVs unique to individual probands, while the second targeted recurrent CNVs overlapping dosage-sensitive regions curated from multiple reference databases. The visualization of candidate CNVs using VizCNV, alongside available phenotypic and familial information, facilitated the manual curation of plausible diagnostic events. Altogether, 36 clinically relevant CNVs were identified. Representative cases, including a *PMP22* duplication consistent with Charcot-Marie-Tooth disease and a complex terminal rearrangement on chromosome 16, highlight the diagnostic contributions of GS-based CNV analysis in families with previously undiagnosed disorders.

In conclusion, this work highlights the outcomes of a community-driven Hackathon focused on advancing CNV/SV detection and interpretation from short-read genome sequencing data. The development and deployment of a shared, queryable CNV/SV database served as a central resource for harmonizing variant interpretation across groups, facilitating reproducibility, transparency, and cross-cohort comparisons. This centralized platform not only enabled standardized variant review but also empowered phenotype-driven exploration and prioritization using clinically relevant annotations.

Together, these efforts underscore the power of community-based collaboration, standardized QC, and shared infrastructure in improving structural variant interpretation. Continued development of such platforms will be key to accelerating rare disease diagnosis and fostering broader insights into the genetic architecture of human disease.

## Data Availability

The GREGoR cohort Data can be accessed at the GREGoR Consortium website. https://gregorconsortium.org/data.

BAM files from PID and TDD participants who consented to share data in public controlled datasets are available in dbGaP phs002999.v5.

## Code Availability

vizCNV: https://github.com/Carvalho-Lab/VizCNV

CNV/SV database: https://github.com/Carvalho-Lab/CLDB

BEDanno: https://github.com/Carvalho-Lab/BEDanno

## Acknowledgments

We thank the families, colleagues and collaborators for their participation in this study. An author of this publication is a member of the European Reference Network on Rare Congenital Malformations and Rare Intellectual Disability ERN-ITHACA [EU Framework Partnership Agreement ID: 3HP-HP-FPA ERN-01-2016/739516].

## Funding Statement

This work was supported by the US National Institute of General Medical Sciences (NIGMS) R01 GM132589 (to CMBC), in part by the National Human Genome Research Institute/National Heart, Lung, and Blood Institute (UM1HG006542, to the Baylor-Hopkins Center for Mendelian Genomics) and the NHGRI Genomic Research Elucidates Genetics of Rare disease (GREGoR) consortium U01 HG011758 to JEP, in part by the Swedish Brain Foundation, grant number FO2022-0256, and the Swedish Rare Diseases Research Foundation (Sällsyntafonden).

## Authors Contributions

Conceptualization: A.L., J.E.P., C.M.B.C.; Formal analysis: M.Y.L., J.E.P., J.D.B., H.D., R.S.R. L.Y., S.O., A.L., C.M.B.C.; Methodology: M.Y.L., H.D., S.O.; Validation: B.Y., M.G.; Writing, review & editing: M.Y.L., J.E.P., S.O., A.L., C.M.B.C; Supervision: J.E.P., C.M.B.C. All authors reviewed the manuscript.

## Ethics Declaration

This study was performed with approval by the Institutional Review Board (IRB) at Baylor College of Medicine and Pacific Northwest Research Institute through WIRB under research protocols H-29697and H-47127/20202158). Informed consent was required by the IRBs.

## Conflict of Interest

Jennifer Posey serves on the Advisory Board of MaddieBio. Other authors declare no conflict of interest.

---

**Supp Figure 1:** PID and Turkish cohort read depth QC results. A) Density plot of standard deviation of log2 ratio for all qualified segments. Red dotted line shows the log2 ratio threshold of 0.38. B) Correlation plot between qualified segments counts and SD value showed negative correlation like the GREGoR samples.

**Supp Figure 2:** Main phenotype groups for the Turkish Developmental Disorder cohort.

**Supp Figure 3:** VizCNV plots for pathogenic CNVs in case BH9683-1. The probands carry large *de novo* chr15q11-13 deletions. The deletion occurred in the paternal chromosome consistent with Prader-Willi syndrome.

**Supp Figure 4:** VizCNV plots for pathogenic CNVs in case BH10684-1. The proband carry large *de novo* chr15q11-13 deletions. The deletion occurred in the maternal chromosome consistent with Angelman syndrome.

**Supp Figure 5:** VizCNV plot for pathogenic CNV in Case BH13688-1. A *de novo* partial deletion in *ZC4H2* is observed. From the read-depth plot, only the proband displays log2 ratio consistent with heterozygous deletion which is supported by the B-allele frequency plot.

**Supp Figure 6:** VizCNV plot for VUS CNV in Case BH13674-1. A large 149kb paternally inherited intronic deletion is observed in chr20 in *MACROD2*. The absence of heterozygosity in the region in the B-allele frequency plot supports the heterozygous deletion.

**Supp Figure 7:** Visualization of two CNVs prioritized for further review from the GREGoR cohort. Pedigree structure and phenotype information as indicated in each panel inset. A) Duplication on chromosome 17 encompassing *PMP22*, with trio B-allele frequency plot supports a maternal inherited CNV. B) *de novo* likely pathogenic terminal chromosome 16 deletion/duplication. B-allele frequency plot supports the heterozygous deletion and duplication and indicates both CNVs were generated in the paternal chromosome 16.

## References

1. Dawood M, Heavner B, Wheeler MM, Ungar RA, LoTempio J, Wiel L, et al. GREGoR: Accelerating Genomics for Rare Diseases. ArXiv. 2024. Epub 20241218.

2. Philippakis AA, Azzariti DR, Beltran S, Brookes AJ, Brownstein CA, Brudno M, et al. The Matchmaker Exchange: a platform for rare disease gene discovery. Hum Mutat. 2015;36:915–921. doi: 10.1002/humu.22858.

3. Deb S, Kalra D, Kubica J, Stricker E, Truong V, Zeng Q, et al. The fifth international hackathon for developing computational cloud-based tools and resources for pan-structural variation and genomics [version 1; peer review: 2 approved with reservations]. F1000Research. 2024;13. doi: 10.12688/f1000research.148237.1.

4. Delgado-Vega AM, Cederroth H, Taylan F, Ekholm K, Ek M, Thonberg H, et al. Pushing the boundaries of rare disease diagnostics with the help of the first Undiagnosed Hackathon. Nat Genet. 2024;56:2287–2294. doi: 10.1038/s41588-024-01941-1.

5. Trzupek K, Bhargava R, Kuan C, Sie F, Vogel-Farley V, Hobbs K, et al. Breaking barriers in rare disease research: The RARE-X Open Science Data Challenge as a model for collaborative innovation and community partnership. HGG Adv. 2025;6:100462. Epub 20250530. doi: 10.1016/j.xhgg.2025.100462.

6. Robinson JT, Thorvaldsdottir H, Winckler W, Guttman M, Lander ES, Getz G, et al. Integrative genomics viewer. Nat Biotechnol. 2011;29:24–26. doi: 10.1038/nbt.1754.

7. Du H, Lun MY, Gagarina L, Mehaffey MG, Hwang JP, Jhangiani SN, et al. VizCNV: An integrated platform for concurrent phased BAF and CNV analysis with trio genome sequencing data. bioRxiv. 2024:2024.2010.2027.620363.doi: 10.1101/2024.10.27.620363.

8. Zarate S, Carroll A, Mahmoud M, Krasheninina O, Jun G, Salerno WJ, et al. Parliament2: Accurate structural variant calling at scale. Gigascience. 2020;9. doi: 10.1093/gigascience/giaa145.

9. Pedersen BS, Quinlan AR. Mosdepth: quick coverage calculation for genomes and exomes. Bioinformatics. 2018;34:867–868. doi: 10.1093/bioinformatics/btx699.

10. Orlandini V, Provenzano A, Giglio S, Magi A. SLMSuite: a suite of algorithms for segmenting genomic profiles. BMC Bioinformatics. 2017;18:321. Epub 20170628. doi: 10.1186/s12859-017-1734-5.

11. Pinto D, Darvishi K, Shi X, Rajan D, Rigler D, Fitzgerald T, et al. Comprehensive assessment of array-based platforms and calling algorithms for detection of copy number variants. Nat Biotechnol. 2011;29:512–520. Epub 20110508. doi: 10.1038/nbt.1852.

12. Ester M, Kriegel H-P, Sander J, Xu X. A density-based algorithm for discovering clusters in large spatial databases with noise. Proceedings of the Second International Conference on Knowledge Discovery and Data Mining; Portland, Oregon: AAAI Press; 1996. p. 226–231.

13. Quinlan AR, Hall IM. BEDTools: a flexible suite of utilities for comparing genomic features. Bioinformatics. 2010;26:841–842. Epub 20100128. doi: 10.1093/bioinformatics/btq033.

14. Geoffroy V, Herenger Y, Kress A, Stoetzel C, Piton A, Dollfus H, et al. AnnotSV: an integrated tool for structural variations annotation. Bioinformatics. 2018;34:3572–3574. doi: 10.1093/bioinformatics/bty304.

15. Pruitt KD, Brown GR, Hiatt SM, Thibaud-Nissen F, Astashyn A, Ermolaeva O, et al. RefSeq: an update on mammalian reference sequences. Nucleic Acids Res. 2014;42:D756–763. Epub 20131119. doi: 10.1093/nar/gkt1114.

16. Bailey JA, Gu Z, Clark RA, Reinert K, Samonte RV, Schwartz S, et al. Recent segmental duplications in the human genome. Science. 2002;297:1003–1007. doi: 10.1126/science.1072047.

17. Rehm HL, Berg JS, Brooks LD, Bustamante CD, Evans JP, Landrum MJ, et al. ClinGen-- the Clinical Genome Resource. N Engl J Med. 2015;372:2235–2242. Epub 20150527. doi: 10.1056/NEJMsr1406261.

18. Collins RL, Glessner JT, Porcu E, Lepamets M, Brandon R, Lauricella C, et al. A cross-disorder dosage sensitivity map of the human genome. Cell. 2022;185:3041–3055 e3025. Epub 20220801. doi: 10.1016/j.cell.2022.06.036.

19. DiStefano MT, Goehringer S, Babb L, Alkuraya FS, Amberger J, Amin M, et al. The Gene Curation Coalition: A global effort to harmonize gene-disease evidence resources. Genet Med. 2022;24:1732–1742. Epub 20220504. doi: 10.1016/j.gim.2022.04.017.

20. Pavan S, Rommel K, Mateo Marquina ME, Hohn S, Lanneau V, Rath A. Clinical Practice Guidelines for Rare Diseases: The Orphanet Database. PLoS One. 2017;12:e0170365. Epub 20170118. doi: 10.1371/journal.pone.0170365.

21. Chen S, Francioli LC, Goodrich JK, Collins RL, Kanai M, Wang Q, et al. A genomic mutational constraint map using variation in 76,156 human genomes. Nature. 2024;625:92–100. Epub 20231206. doi: 10.1038/s41586-023-06045-0.

22. Amberger J, Bocchini CA, Scott AF, Hamosh A. McKusick’s Online Mendelian Inheritance in Man (OMIM). Nucleic Acids Res. 2009;37:D793–796. Epub 20081008. doi: 10.1093/nar/gkn665.

23. Tucci V, Isles AR, Kelsey G, Ferguson-Smith AC, Erice Imprinting G. Genomic Imprinting and Physiological Processes in Mammals. Cell. 2019;176:952–965. doi: 10.1016/j.cell.2019.01.043.

24. Fernandez-Luna L, Aguilar-Perez C, Grochowski CM, Mehaffey MG, Carvalho CMB, Gonzaga-Jauregui C. Genome-wide maps of highly-similar intrachromosomal repeats that can mediate ectopic recombination in three human genome assemblies. HGG Adv. 2025;6:100396. Epub 20241224. doi: 10.1016/j.xhgg.2024.100396.

25. Karczewski KJ, Francioli LC, Tiao G, Cummings BB, Alfoldi J, Wang Q, et al. The mutational constraint spectrum quantified from variation in 141,456 humans. Nature. 2020;581:434–443. Epub 20200527. doi: 10.1038/s41586-020-2308-7.

26. Baxter SM, Posey JE, Lake NJ, Sobreira N, Chong JX, Buyske S, et al. Centers for Mendelian Genomics: A decade of facilitating gene discovery. Genet Med. 2022;24:784–797. Epub 20220209. doi: 10.1016/j.gim.2021.12.005.

27. Bousfiha A, Moundir A, Tangye SG, Picard C, Jeddane L, Al-Herz W, et al. The 2022 Update of IUIS Phenotypical Classification for Human Inborn Errors of Immunity. J Clin Immunol. 2022;42:1508–1520. Epub 20221006. doi: 10.1007/s10875-022-01352-z.

28. Renner ED, Puck JM, Holland SM, Schmitt M, Weiss M, Frosch M, et al. Autosomal recessive hyperimmunoglobulin E syndrome: a distinct disease entity. J Pediatr. 2004;144:93–99. doi: 10.1016/S0022-3476(03)00449-9.

29. Coban-Akdemir Z, Song X, Ceballos FC, Pehlivan D, Karaca E, Bayram Y, et al. The impact of the Turkish population variome on the genomic architecture of rare disease traits. Genet Med Open. 2024;2:101830. Epub 20240214. doi: 10.1016/j.gimo.2024.101830.

30. Buiting K, Saitoh S, Gross S, Dittrich B, Schwartz S, Nicholls RD, et al. Inherited microdeletions in the Angelman and Prader-Willi syndromes define an imprinting centre on human chromosome 15. Nat Genet. 1995;9:395–400. doi: 10.1038/ng0495-395.

31. Cassidy SB, Lai LW, Erickson RP, Magnuson L, Thomas E, Gendron R, et al. Trisomy 15 with loss of the paternal 15 as a cause of Prader-Willi syndrome due to maternal disomy. Am J Hum Genet. 1992;51:701–708.

32. Frints SGM, Hennig F, Colombo R, Jacquemont S, Terhal P, Zimmerman HH, et al. Deleterious de novo variants of X-linked ZC4H2 in females cause a variable phenotype with neurogenic arthrogryposis multiplex congenita. Hum Mutat. 2019;40:2270–2285. Epub 20190821. doi: 10.1002/humu.23841.

33. Sun JJ, Cai Q, Xu M, Liu YN, Li WR, Li J, et al. Loss of Protein Function Causing Severe Phenotypes of Female-Restricted Wieacker Wolff Syndrome due to a Novel Nonsense Mutation in the ZC4H2 Gene. Genes (Basel). 2022;13. Epub 20220829. doi: 10.3390/genes13091558.

34. Lombardo B, Esposito D, Iossa S, Vitale A, Verdesca F, Perrotta C, et al. Intragenic Deletion in *MACROD2*: A Family with Complex Phenotypes Including Microcephaly, Intellectual Disability, Polydactyly, Renal and Pancreatic Malformations. Cytogenet Genome Res. 2019;158:25–31. Epub 20190503. doi: 10.1159/000499886.

35. Roa BB, Garcia CA, Lupski JR. Charcot-Marie-Tooth disease type 1A: molecular mechanisms of gene dosage and point mutation underlying a common inherited peripheral neuropathy. Int J Neurol. 1991;25-26:97–107.

36. Hermetz KE, Newman S, Conneely KN, Martin CL, Ballif BC, Shaffer LG, et al. Large inverted duplications in the human genome form via a fold-back mechanism. PLoS Genet. 2014;10:e1004139. Epub 20140130. doi: 10.1371/journal.pgen.1004139.

37. Zhang Q, Davis JC, Lamborn IT, Freeman AF, Jing H, Favreau AJ, et al. Combined immunodeficiency associated with *DOCK8* mutations. N Engl J Med. 2009;361:2046–2055. Epub 20090923. doi: 10.1056/NEJMoa0905506.

