## Supplementary material for "Community-Driven Copy Number Variant Discovery at Scale: Results from a Rare Disease Genomics Hackathon": Supp Figure

### Slide 1
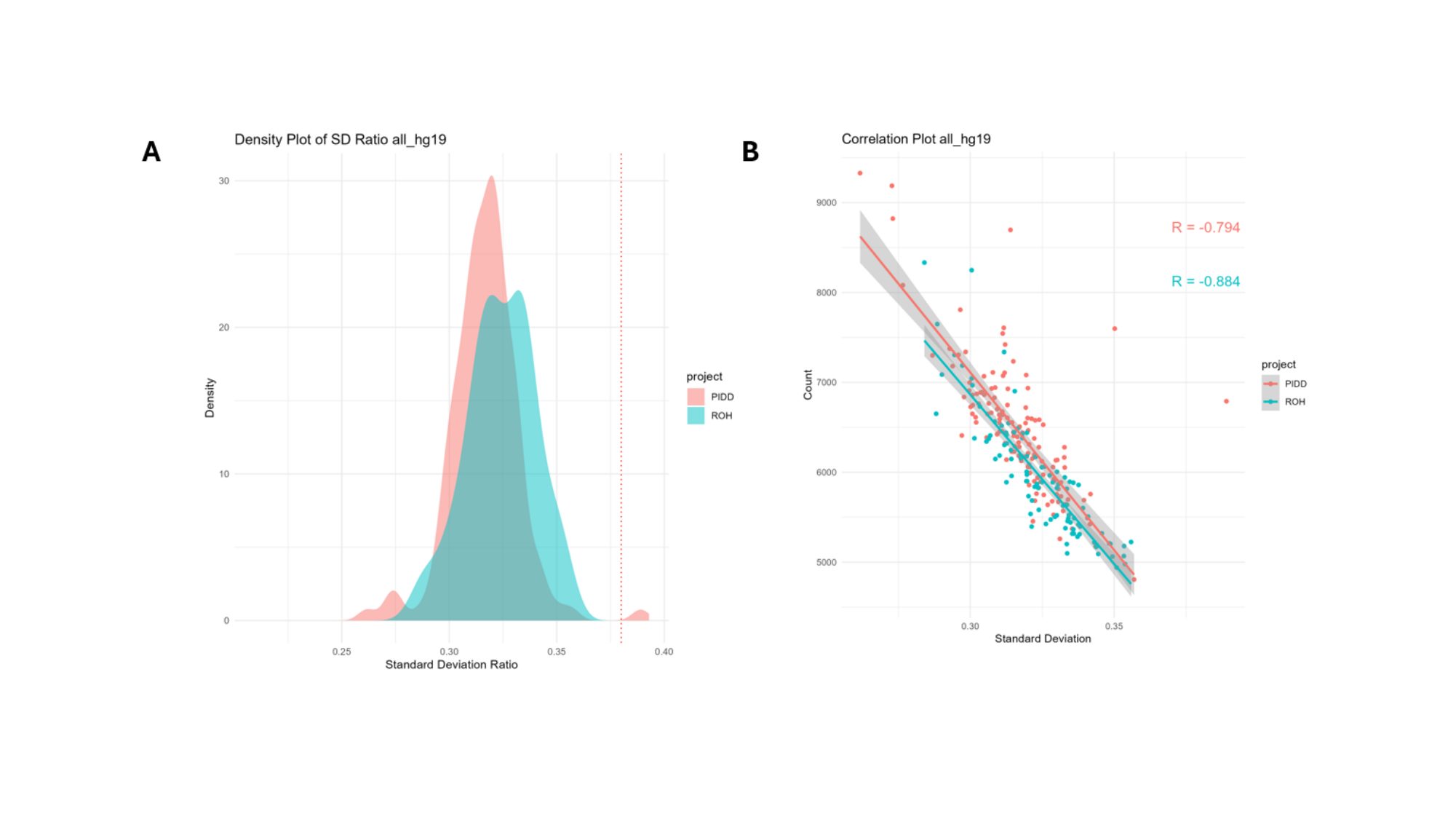

### Slide 2
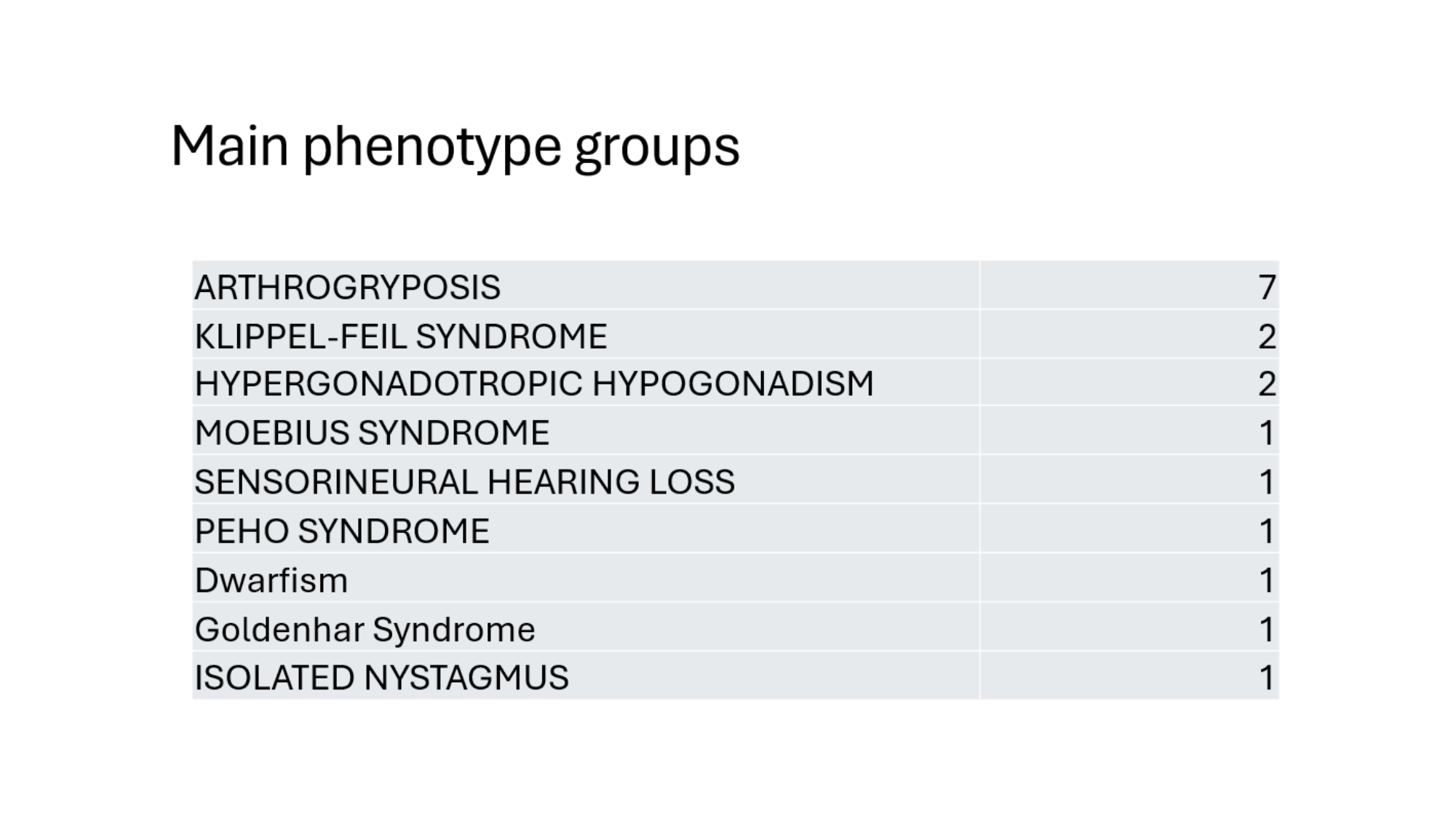

### Slide 3
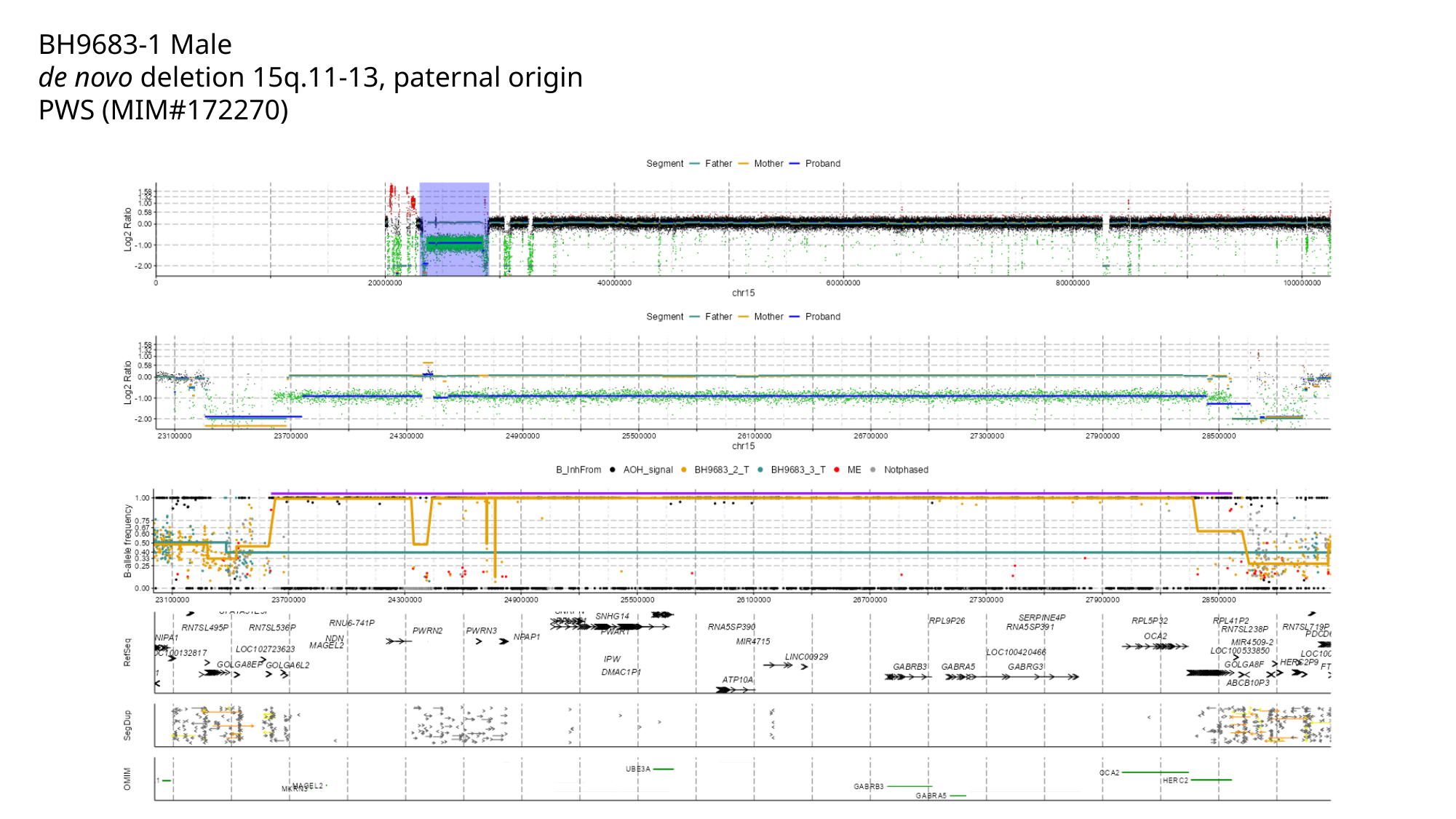

BH9683-1 Male
de novo deletion 15q.11-13, paternal origin
PWS (MIM#172270)

### Slide 4
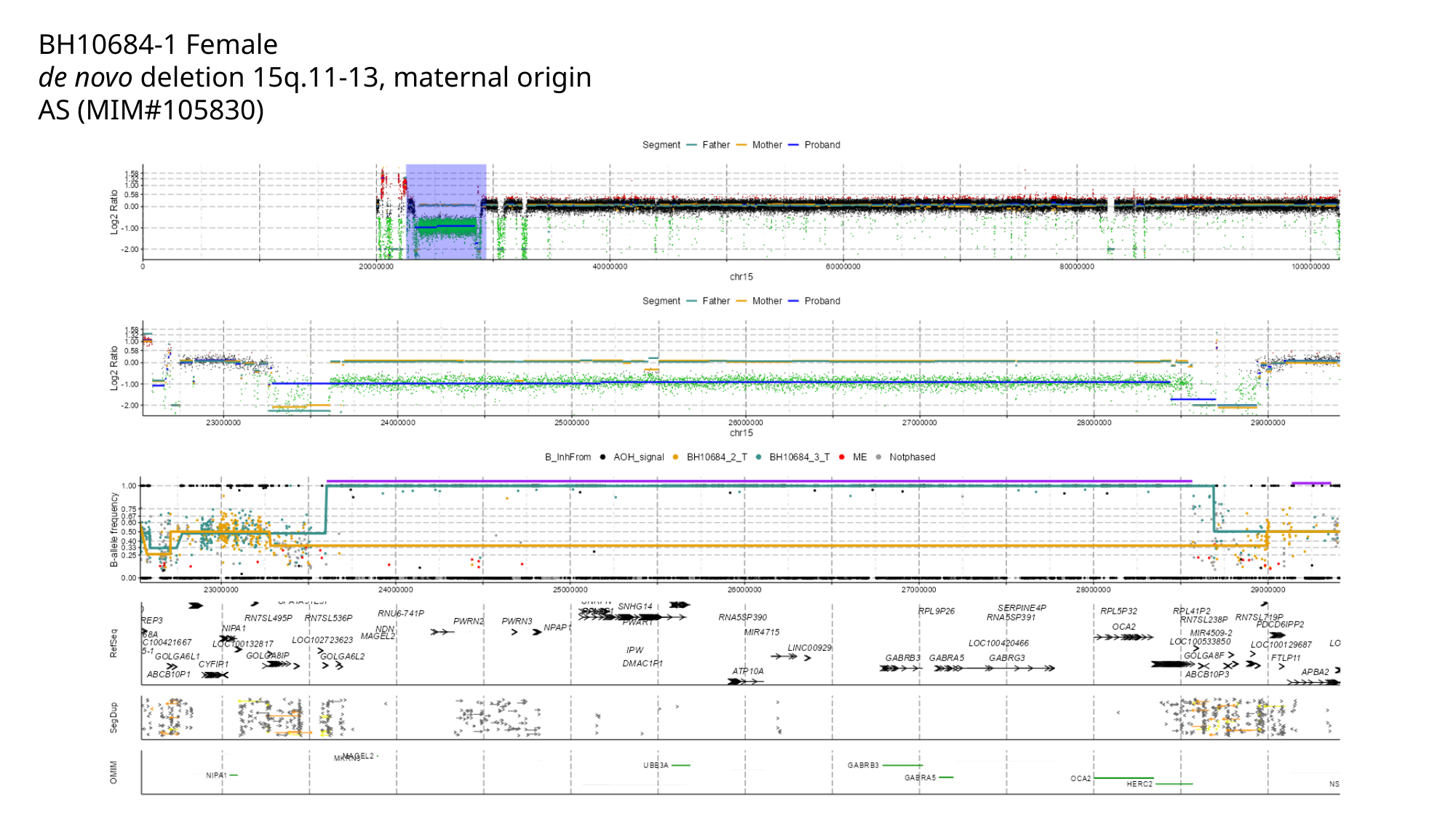

BH10684-1 Female
de novo deletion 15q.11-13, maternal origin
AS (MIM#105830)

### Slide 5
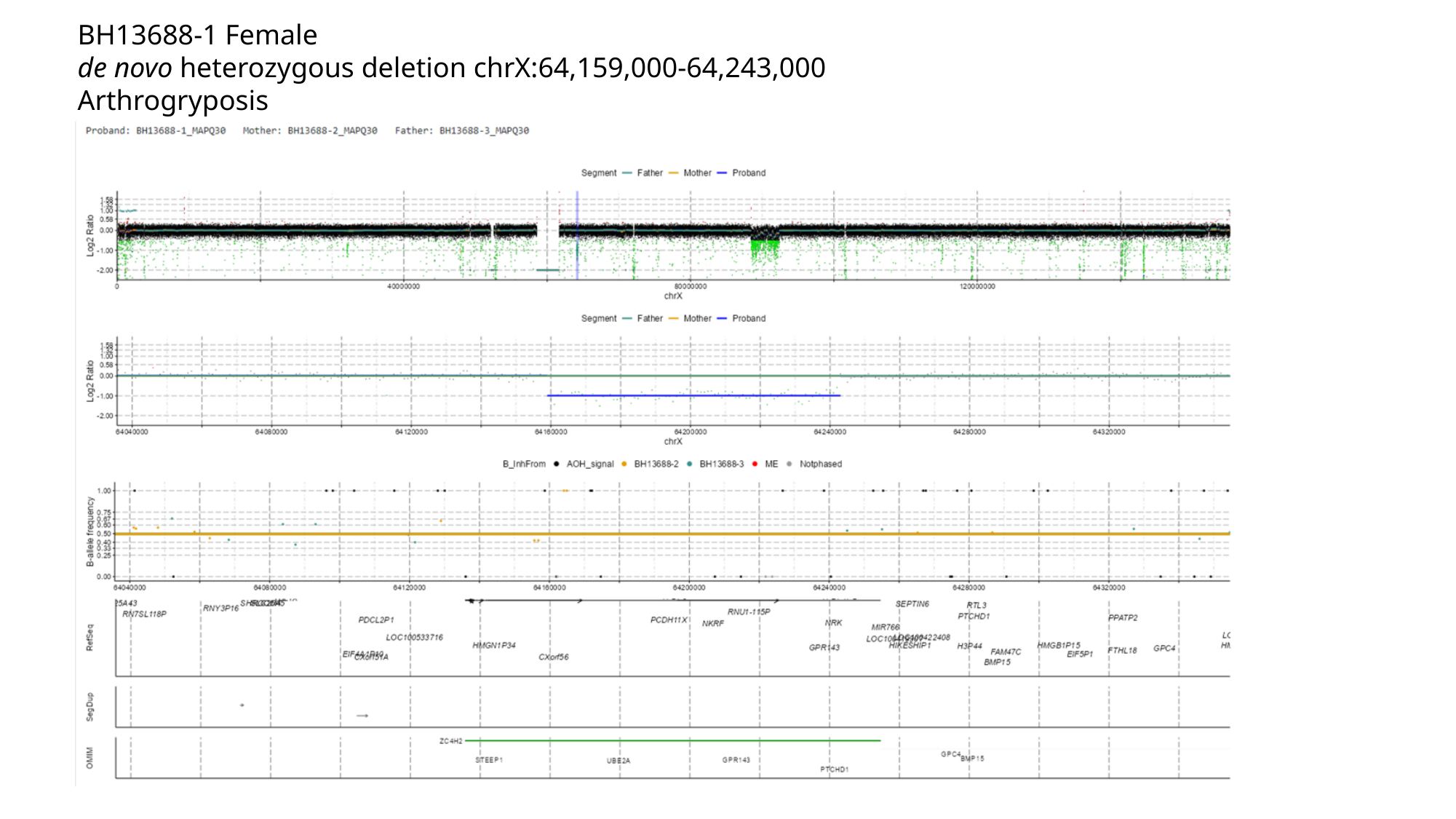

BH13688-1 Female
de novo heterozygous deletion chrX:64,159,000-64,243,000
Arthrogryposis
#

### Slide 6
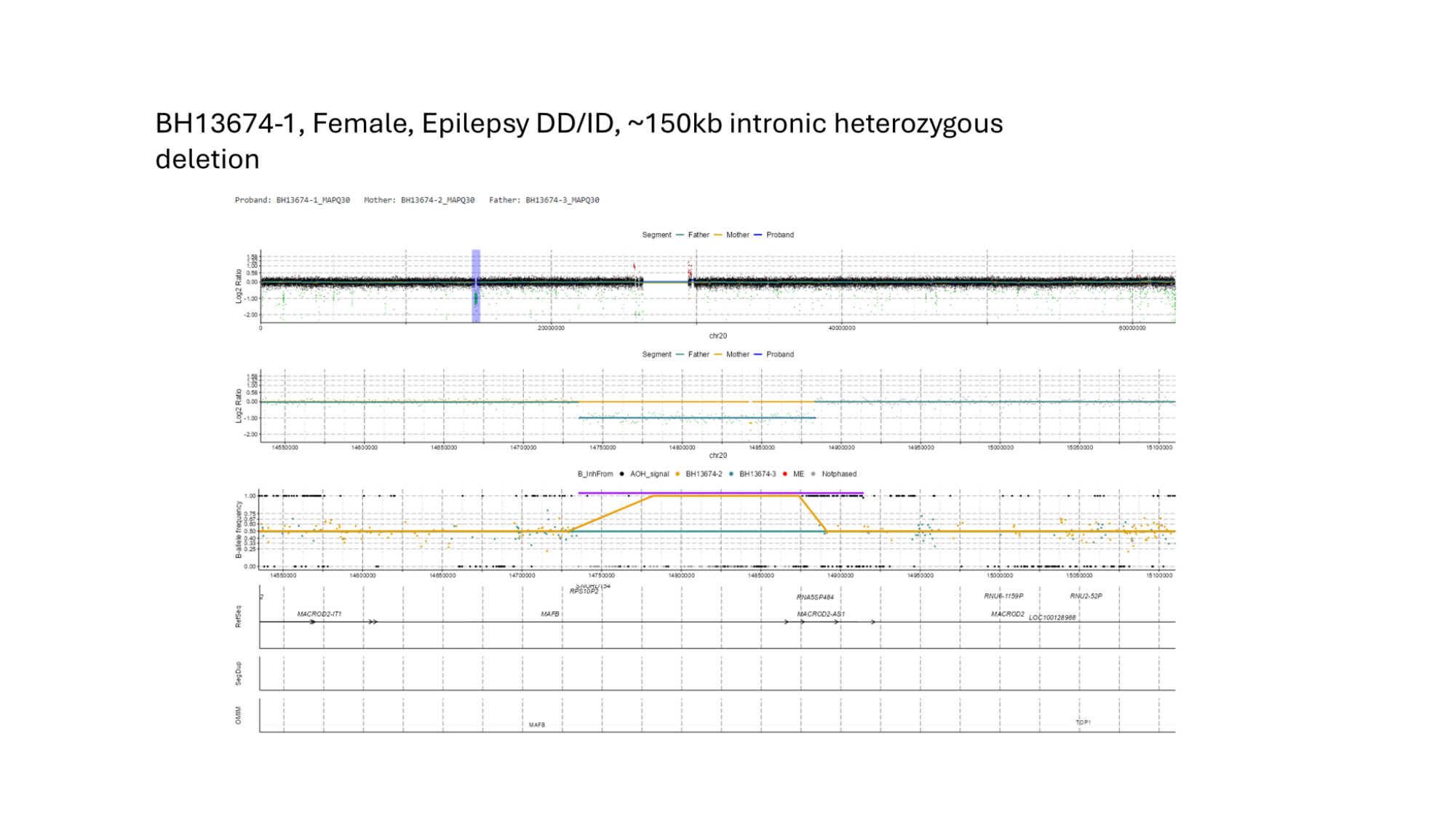

### Slide 7
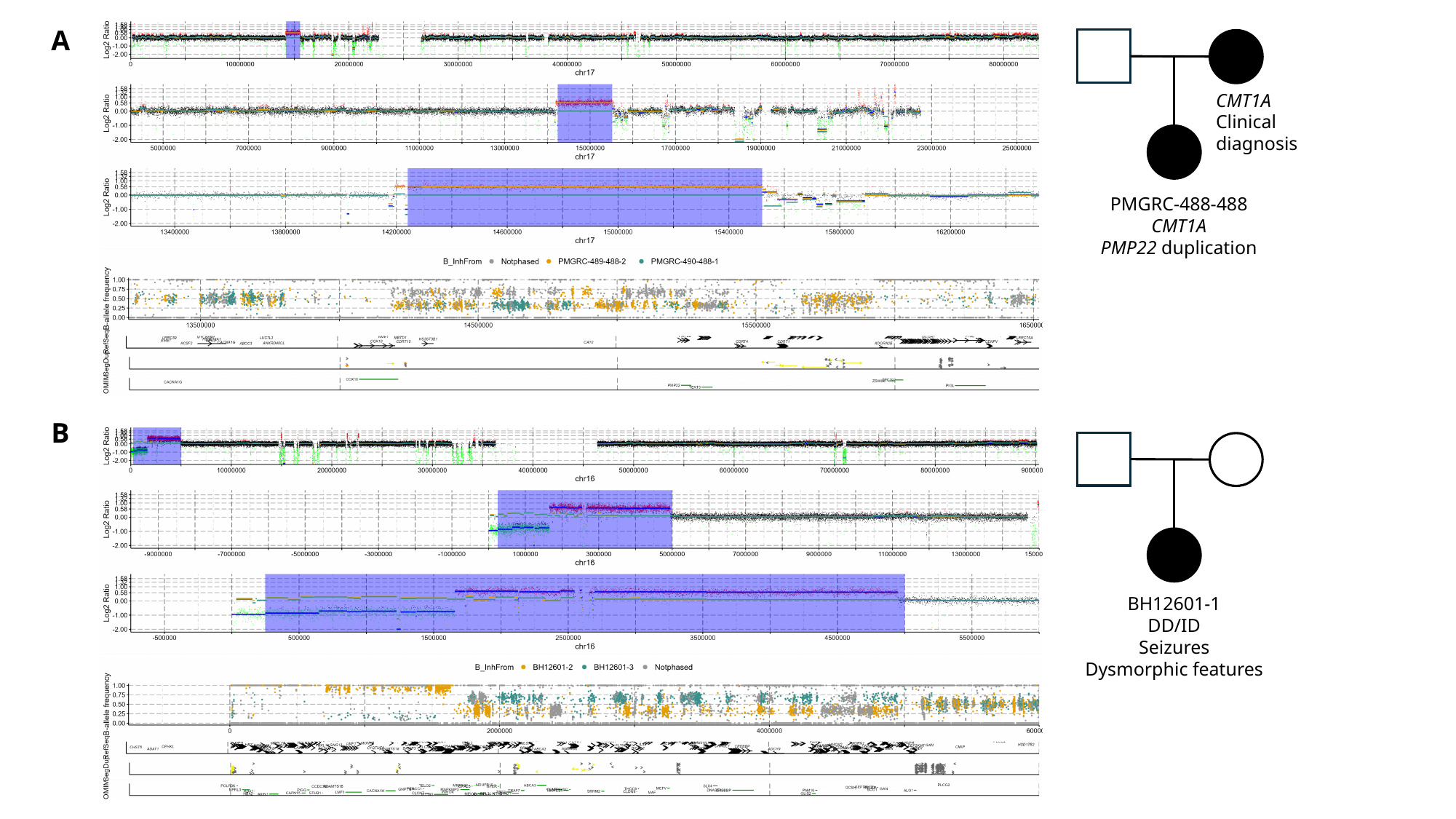

A
PMGRC-488-488
CMT1A
PMP22 duplication
CMT1A
Clinical diagnosis
B
BH12601-1
DD/ID
Seizures
Dysmorphic features
